# Effectiveness of the Single-Dose Ad26.COV2.S COVID Vaccine

**DOI:** 10.1101/2021.09.10.21263385

**Authors:** Jennifer M. Polinski, Andrew R. Weckstein, Michael Batech, Carly Kabelac, Tripthi Kamath, Raymond Harvey, Sid Jain, Jeremy A. Rassen, Najat Khan, Sebastian Schneeweiss

## Abstract

**Importance:** Vaccination against the SARS-CoV-2 virus is critical to control the pandemic. Randomized trials demonstrated efficacy of the single-dose Ad26.COV2.S COVID vaccine but data on longer-term protection in clinical practice and effectiveness against variants are needed.

**Objective:** To assess the effectiveness of Ad26.COV2.S in preventing COVID infections and COVID-related hospitalizations in clinical practice, the longer-term stability of its protective effect and effectiveness against Delta variants.

**Design:** Cohort study of newly Ad26.COV2.S-vaccinated and unvaccinated individuals.

**Setting:** U.S. insurance claims data through July 2021.

**Participants:** Individuals 18 years and older newly vaccinated with Ad26.COV2.S and up to 10 unvaccinated individuals matched exactly by age, sex, date, location, comorbidity index plus 17 COVID-19 risk factors via propensity score (PS) matching.

**Intervention:** Vaccination with Ad26.COV2.S versus no vaccination.

**Main outcomes:** We estimated vaccine effectiveness (VE) for observed COVID-19 infection and COVID-19-related hospitalization, nationwide and stratified by age, immunocompromised status, calendar time, and states with high incidence of the Delta variant. We corrected VE estimates for under-recording of vaccinations in insurance data.

**Results:** Among 390,517 vaccinated and 1,524,153 matched unvaccinated individuals, VE was 79% (95% CI, 77% to 80%) for COVID-19 and 81% (79% to 84%) for COVID-19-related hospitalizations. VE was stable over calendar time. Among states with high Delta variant incidence, VE during June/July 2021 was 78% (73% to 82%) for infections and 85% (73% to 91%) for hospitalizations. VE for COVID-19 was higher in individuals <50 years (83%; 81% to 85%) and lower in immunocompromised patients (64%; 57% to 70%). All estimates were corrected for under-recording; uncorrected VE was 69% (67% to 71%) and 73% (69% to 76%), for COVID-19 and COVID-19-related hospitalization, respectively.

**Conclusions:** These non-randomized data across U.S. clinical practices show high and stable vaccine effectiveness of Ad26.COV2.S over time before the Delta variant emerged to when the Delta variant was dominant.

## Introduction

With the global morbidity and mortality associated with the SARS-CoV-2 virus and the rapid spread of new variants, a temporal and geographical understanding of vaccine effectiveness (VE) in clinical practice is critical to inform public health recommendations. On February 27, 2021, the Food and Drug Administration issued an Emergency Use Authorization for Janssen’s single dose COVID-19 vaccine, Ad26.COV2.S,^1^ based on results from a double-blind, placebo-controlled randomized controlled trial (RCT), ENSEMBLE.^2^ The overall efficacy estimate for preventing moderate-severe observed COVID-19 at least 14 days post-vaccination was 66% globally (74% US) and overall efficacy at least 28 days after vaccination was >84% against severe disease.^3^

Despite the efficacy observed in the RCT, questions remain about generalizability to patients in clinical practice, and longer-term effectiveness. VE may be affected by disease variants, such as the Delta (B.1.617.2, AY.1, AY.2, AY.3) which emerged in late May 2021 and soon became the dominant variant across the U.S. by July.^4^ A recent study showed the effectiveness of Ad26.COV2.S and other COVID-19 vaccines in ambulatory and inpatient care settings using data through June 22, 2021.^5^ However, the study included less than 1,500 subjects who had received Ad26.COV2.S vaccination which limits the interpretation of findings, its generalizability and any statements on the protection against Delta variants, in addition to general drawbacks of the test-negative study design.^6^

As recommendations and guidelines change in response to a rapidly-changing disease landscape, population-based healthcare databases may observe VE, longer-term effectiveness, and COVID-related health outcomes with little delay. We utilized national U.S. claims data to assess the VE of Ad26.COV2.S in preventing breakthrough COVID-19 infections or COVID-19-related hospitalizations from March 2021 through July 2021. We specifically assessed VE during June and July in four U.S. states with early and high prevalence of the Delta variant.^7^

## Methods

### Data

We analyzed de-identified patient-level claims data from March 1, 2020 through July 31, 2021 submitted to insurance companies by U.S. providers of inpatient, outpatient, pharmacy, and laboratory services, and aggregated by HealthVerity.^8^ The claims were anonymously linked across >70 different health data sources using a unique, encrypted and non-identifiable patient token derived from identifiable information known only to the provider. This token allows for cross-database longitudinal capture of diagnosis and procedure codes, COVID-19 laboratory test orders and results, and pharmacy dispensing information.^9^ Cohorts for the current study were identified from a larger dataset of 160 million individuals with any record of lab orders, diagnoses, or treatments broadly related to COVID (COVID-related utilization), or COVID vaccinations from December 2019 to July 2021.

This study was approved by the New England institutional review board. These data have been used in previous studies.^10–12^ They belong to third parties and cannot be shared publicly, however, upon reasonable request, researchers may get access to the data and analytics infrastructure for prespecified collaborative analyses.

### Population and Exposure

Individuals 18 or older who received a single dose of Ad26.COV2.S between March 1, 2021 and July 17, 2021 entered the study cohort on the day of vaccination. Each participant was matched on the same day to up to 10 referent individuals with no evidence of vaccination with a COVID-19 vaccine at that time, with matching by location (3-digit ZIP), age within 4 years, sex, and general health status captured in a comorbidity score (**Suppl. S1**). ^13^ We required at least one medical and pharmacy claim during 365 days before cohort entry to ensure each individual’s activity in the system. Those with observed COVID-19 or receipt of any COVID-19 vaccine during the 365 days before cohort entry were excluded (**Suppl. S2**).

### Follow-up and Outcomes

Follow-up in each group started 14 days after cohort entry^14^ and continued until the earliest of the occurrence of an outcome, receipt of any COVID-19 vaccine, death, or July 31, 2021.

The outcome of observed COVID-19 was defined by either recording of an in- or outpatient ICD-10-CM diagnosis code of U07.1 (85% of cases) in any position, and/or a recorded positive SARS-CoV-2 diagnostic PCR or nucleic acid amplification test result (15%). The co-primary outcome was COVID-19-related hospitalization, defined as any claim for an inpatient stay with a discharge diagnosis of COVID-19 or a recorded infection within 21 days before admission.

### Participant characteristics

Participant characteristics, including a range of factors associated with severe COVID-19 disease,^15^ were assessed during the 365 days before cohort entry (**Suppl. S3** for all definitions): chronic obstructive pulmonary disease, asthma, pulmonary fibrosis, cystic fibrosis, HIV, immunocompromised status after transplantation, liver disease, malignancies, cerebrovascular disease, chronic kidney disease, hypertension, obesity, heart failure, sickle-cell disease, thalassemia, diabetes mellitus, chronic neurologic conditions, and an overall comorbidity score.^13^ We measured healthcare utilization intensity, a marker of patients’ baseline health state and surveillance, including the number of pharmacy or medical claims in 365 days before cohort entry, and recent medical or pharmacy claims in the 60 days before.

### Statistical analysis

To ensure balance between treatment groups in the absence of baseline randomization, and in addition to the 1:10 risk-set sampling by time, location, age, sex, and comorbidity score described above, we further matched the risk-set sampled population using propensity scores (PS). Each vaccinated individual was PS-matched with up to 4 unvaccinated individuals with a caliper of ±0.01. Propensity scores were estimated with logistic regression including all pre-exposure patient characteristics with no further variable selection. The resulting balance of characteristics between the vaccinated and PS-matched unvaccinated individuals was assessed by computing the absolute standardized difference for each covariate.^16^ A value <0.10 indicates little imbalance between treatment groups and little effect on residual confounding.^17^

We computed incidence rates per 1,000 person-years for vaccinated and unvaccinated participants and hazard ratios (HRs) with 95% confidence intervals (CI).^18^ VE in percent was computed as (1-HR)×100. Kaplan-Meier curves with 95% CIs and Schoenfeld residuals were plotted to test whether VE changes over time since vaccination.^19^

Additional analyses included stratification by calendar months March 2021 through July 2021 to determine whether there was a consistent vaccine effect over calendar time (**Suppl. S5**), stratification by age (<50 and ≥50 years; <60 and ≥60), stratification by U.S. region (West, Northeast, Midwest, South), and an analysis of immunocompromised individuals. To examine VE in a time and region where a large fraction of new cases were patients infected with the Delta variant, we separately analyzed data from four states with highest COVID-19 incidence and high proportion of the Delta variant in June and July (Florida, Louisiana, Arkansas, and Missouri, hereafter “high-Delta-incidence states”).^20^

Given the expedited national vaccination effort, a sizable proportion of COVID-19 vaccinations were administered by employers, mass vaccination sites, pharmacies, and other settings where often no health insurance claims were submitted. The CDC reported that 57% of US residents 12 years and older were vaccinated as of July 22, 2021, while only 34% were recorded among the same people in our claims data, which confirms substantial under-recording (**Suppl. S4**). As a result, it is highly likely that a substantial proportion of the unvaccinated group in claims data was in fact vaccinated and thus observed VE estimates will appear lower than indeed true. To compensate, we conservatively assumed 40% under-recording of vaccinations and applied a correction factor to all VE estimates using standard methods for correcting exposure misclassification. In sensitivity analyses, we varied the under-recording assumption from 0% to 70% (**Tables S5 and S6**). ^21^

We conducted two additional sensitivity analyses to explore potential bias related to outcome capture and population sampling. First, we narrowed COVID-19 outcome definitions to require a positive nucleic acid amplification test result (**Table S7**). Second, to minimize potential bias related to non-differential sampling of vaccinated and unvaccinated individuals at the data extraction stage, we required all individuals to have evidence of potentially COVID-related utilization before cohort entry (**Table S8**).

All analyses were performed using the Aetion Evidence Platform v4.13 (including R v3.4.2), which has been scientifically validated by accurately repeating a range of previously-published studies^22^ and by replicating^23^ or predicting clinical trial findings.^24^ All transformations of the raw data are preserved for full reproducibility and audit trails are available, including a quality check of the data ingestion process.

## Results

The study cohort included a total of 390,517 vaccinated U.S. individuals and 1,524,153 matched individuals with no recording of vaccination from March 1, 2021 through July 17, 2021 (**Figure 1**). The two-stage matching by time, location, socio-demographic variables, and risk factors resulted in highly similar groups with absolute standardized mean differences ≤0.02 across all characteristics, well below 0.10 (**Table 1**). In high-Delta-incidence states, 29,079 vaccinated individuals were similarly well-balanced when compared to 113,325 unvaccinated individuals, with all absolute standardized mean differences ≤0.03 (**Table S2**), well below the 0.10 threshold.

**Table 1.** Characteristics of Ad26.COV2.S vaccinated and matched unvaccinated individuals in the national cohort.

| *n (%) or mean +/- SD unless otherwise noted* | **Vaccinated group**<br>**(Exposed)**<br>**n=390,517** | **Unvaccinated group**<br>**(Referent)**<br>**n=1,524,153** | **Absolute**<br>**standardized**<br>**difference** |
| --- | --- | --- | --- |
| Age, mean (sd) | 55.05 (17.31) | 54.94 (17.42) | 0.006 |
| Female sex; n (%) | 219,989 (56.3%) | 858,322 (56.3%) | 0.000 |
| Cerebrovascular disease; n (%) | 14,150 (3.6%) | 55,693 (3.7%) | 0.002 |
| Chronic kidney disease; n (%) | 20,421 (5.2%) | 78,451 (5.1%) | 0.004 |
| Chronic obstructive pulmonary disease; n (%) | 40,375 (10.3%) | 159,065 (10.4%) | 0.003 |
| Cystic fibrosis*; n (%) | 23 (0.0%) | 95 (0.0%) | 0.000 |
| HIV; n (%) | 1,244 (0.3%) | 5,473 (0.4%) | 0.007 |
| Hypertension; n (%) | 120,810 (30.9%) | 470,438 (30.9%) | 0.002 |
| Immunocompromised from blood transplant*; n (%) | 3 (0.0%) | 28 (0.0%) | 0.003 |
| Immunocompromised from organ transplant; n (%) | 1,433 (0.4%) | 6,048 (0.4%) | 0.005 |
| Liver disease; n (%) | 15,910 (4.1%) | 61,785 (4.1%) | 0.001 |
| Malignancies; n (%) | 17,434 (4.5%) | 68,418 (4.5%) | 0.001 |
| Moderate-to-severe asthma; n (%) | 3,504 (0.9%) | 14,214 (0.9%) | 0.004 |
| Neurologic conditions; n (%) | 110,394 (28.3%) | 431,470 (28.3%) | 0.001 |
| Obesity; n (%) | 58,856 (15.1%) | 228,861 (15.0%) | 0.002 |
| Pulmonary fibrosis; n (%) | 1,969 (0.5%) | 8,268 (0.5%) | 0.005 |
| Serious heart conditions; n (%) | 38,895 (10.0%) | 153,329 (10.1%) | 0.003 |
| Sickle-cell disease; n (%) | 160 (0.0%) | 754 (0.0%) | 0.004 |
| Thalassemia; n (%) | 190 (0.0%) | 768 (0.1%) | 0.001 |
| Type 1 diabetes; n (%) | 4,120 (1.1%) | 16,135 (1.1%) | 0.000 |
| Type 2 diabetes; n (%) | 60,410 (15.5%) | 233,175 (15.3%) | 0.005 |
| Gagne combined comorbidity score, mean (sd) | 0.64 (1.56) | 0.63 (1.55) | 0.003 |
| Count of medical claims | 12.76 (33.33) | 12.91 (31.17) | 0.005 |
| Count of pharmacy claims | 17.97 (18.88) | 17.83 (18.50) | 0.008 |
| Recent medical claim** | 192,474 (49.3%) | 749,810 (49.2%) | 0.002 |
| Recent pharmacy claim** | 277,989 (71.2%) | 1,077,266 (70.7%) | 0.011 |
| Index months |  |  | 0.006 |
| March 2021; n (%) | 146,576 (37.5%) | 572,459 (37.6%) | - |
| April 2021; n (%) | 144,449 (37.0%) | 566,750 (37.2%) | - |
| May 2021; n (%) | 68,082 (17.4%) | 264,268 (17.3%) | - |
| June 2021; n (%) | 25,020 (6.4%) | 96,188 (6.3%) | - |
| July 2021; n (%) | 6,390 (1.6%) | 24,488 (1.6%) | - |
| U.S. Region |  |  | 0.002 |
| Northeast; n (%) | 53,406 (13.7%) | 208,572 (13.7%) | - |
| Midwest; n (%) | 87,045 (22.3%) | 338,650 (22.2%) | - |
| South; n (%) | 162,865 (41.7%) | 636,225 (41.7%) | - |
| West; n (%) | 87,201 (22.3%) | 340,706 (22.4%) | - |
| State |  |  | 0.011 |

Characteristics reported for population matched with risk-set sampling and propensity scores. Unless otherwise noted, demographic variables are assessed at cohort entry (index) and comorbidities and clinical utilization variables are assessed during the 1 year before cohort entry.
\*Variables not included in propensity score models.
\*\*Recent medical and pharmacy claims were defined as claims beginning during the 60 days before cohort entry.

**Figure 1.**
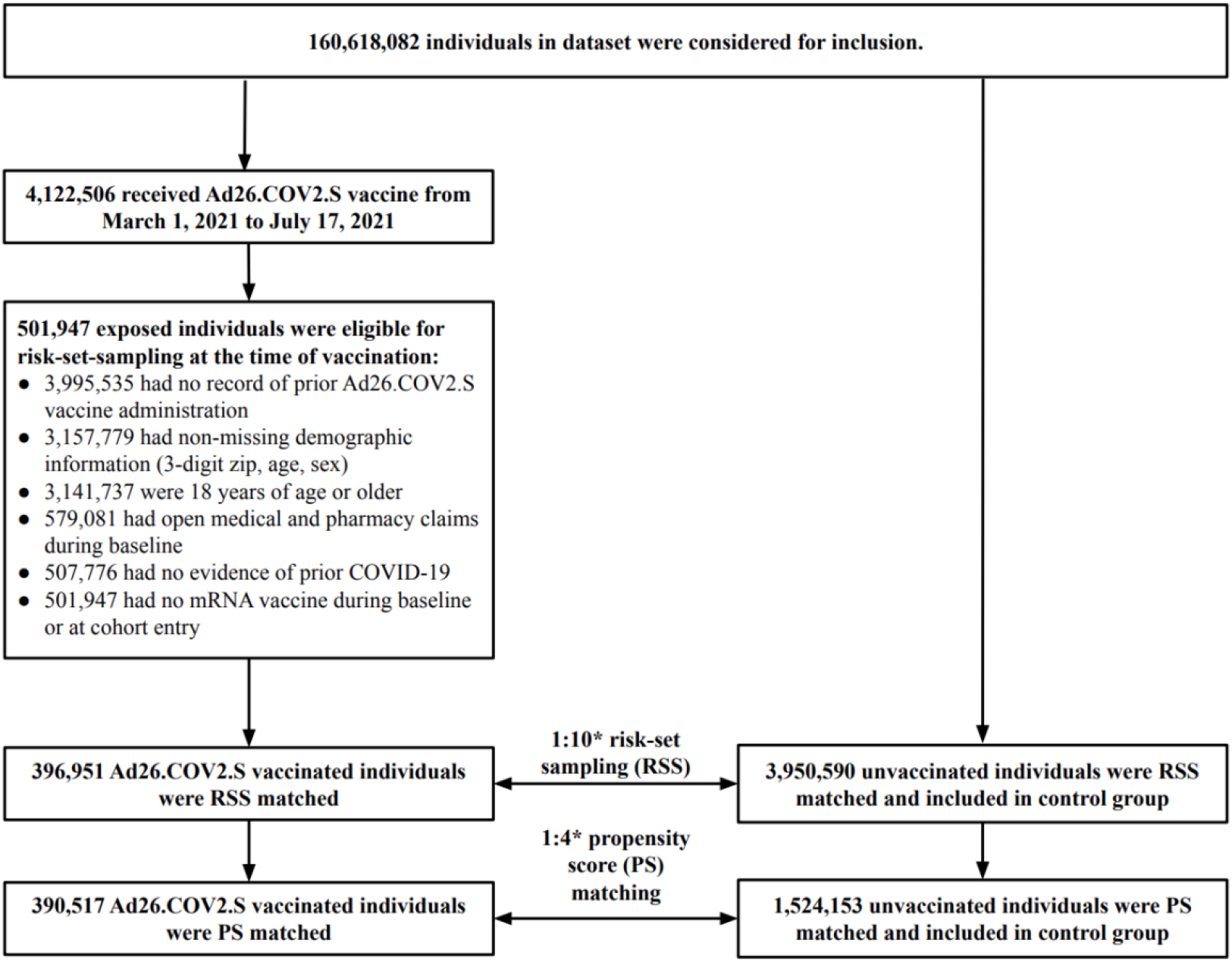
Study population CONSORT diagram. * Match ratios reflect the maximum number of unvaccinated matches allowed for each vaccinated individual, however, some exposed individuals will have fewer than the maximum number of matches. During risk-set sampling, each vaccinated individual was RSS matched with *up to* 10 unvaccinated individuals; for propensity score matching, each vaccinated individual was PS matched with *up to* 4 unvaccinated individuals.

Subgroups defined by age category, immunocompromised status, and geography were similarly well balanced (**Tables S1 and S2**). A majority of event-free individuals reached the end of the study period (99.3% of vaccinated and 81.9% of matched unvaccinated), with 18.1% of unvaccinated individuals censored for receipt of COVID-19 vaccine. In the national cohort, 1,232 vaccinated (12.0/1,000 person-years) and 13,505 unvaccinated individuals (39.5/1,000 PY) had an observed COVID-19 (**Table 2**). VE for observed COVID-19 was 79% (95% CI, 77% to 80%). Incidence rates of COVID-19-related hospitalization were 2.4/1,000 PY in vaccinated and 9.1/1,000 PY in unvaccinated individuals resulting in a corrected VE of 81% (79% to 84%). VE for observed COVID-19 was higher in patients <50 years of age (83%; 81% to 85%); than in patients ≥50 (75%; 74% to 77%). Immunocompromised individuals had lower VE (64%; 57% to 70%) compared to non-immunocompromised individuals (79%; 78% to 81%). There was some regional variation in VE (**Table S4**).

**Table 2.** Incidence and vaccine effectiveness for COVID-19 and COVID-19-related hospitalizations, nationally and by subgroup.

|  | Vaccinated group |  |  | Unvaccinated group |  |  | Hazard Ratios<br>(95% CI) | Vaccine<br>Effectiveness<br>(95% CI) | Vaccine Effectiveness<br>(95% CI)<br>Corrected for vaccine<br>under-recording |
| --- | --- | --- | --- | --- | --- | --- | --- | --- | --- |
|  | N<br>events | Person-years | Incidence<br>rate | N events | Person-years | Incidence<br>rate |  |  |  |
| **<u>National cohort</u>** |  |  |  |  |  |  |  |  |  |
| Observed COVID-19 | 1,232 | 102,864 | 11.98 | 13,505 | 341,511 | 39.54 | 0.31 (0.29, 0.33) | 69% (67%, 71%) | 79% (77%, 80%) |
| COVID-19-related hospitalization | 243 | 103,023 | 2.36 | 3,119 | 343,153 | 9.09 | 0.27 (0.24, 0.31) | 73% (69%, 76%) | 81% (79%, 84%) |
| **<u>Subgroups</u>** |  |  |  |  |  |  |  |  |  |
| **Age < 50** |  |  |  |  |  |  |  |  |  |
| Observed COVID-19 | 381 | 32,913 | 11.58 | 5,093 | 108,603 | 46.90 | 0.25 (0.23, 0.28) | 75% (72%, 77%) | 83% (81%, 85%) |
| COVID-19-related hospitalization | 32 | 32,965 | 0.97 | 528 | 109,282 | 4.83 | 0.21 (0.15, 0.30) | 79% (70%, 85%) | 86% (80%, 90%) |
| **Age >= 50** |  |  |  |  |  |  |  |  |  |
| Observed COVID-19 | 851 | 69,951 | 12.17 | 8,368 | 232,614 | 35.97 | 0.35 (0.32, 0.37) | 65% (63%, 68%) | 75% (74%, 77%) |
| COVID-19-related hospitalization | 211 | 70,058 | 3.01 | 2,475 | 233,572 | 10.60 | 0.29 (0.26, 0.34) | 71% (66%, 74%) | 80% (77%, 82%) |
| **Age < 60** |  |  |  |  |  |  |  |  |  |
| Observed COVID-19 | 677 | 55,400 | 12.22 | 7,986 | 181,315 | 44.04 | 0.28 (0.26, 0.31) | 72% (69%, 74%) | 81% (79%, 82%) |
| COVID-19-related hospitalization | 71 | 55,494 | 1.28 | 1,180 | 182,352 | 6.47 | 0.21 (0.16, 0.26) | 79% (74%, 84%) | 86% (83%, 89%) |
| **Age >= 60** |  |  |  |  |  |  |  |  |  |
| Observed COVID-19 | 555 | 47,463 | 11.69 | 5,430 | 160,049 | 33.93 | 0.35 (0.32, 0.39) | 65% (61%, 68%) | 75% (73%, 78%) |
| COVID-19-related hospitalization | 172 | 47,529 | 3.62 | 1,904 | 160,631 | 11.85 | 0.32 (0.27, 0.37) | 68% (63%, 73%) | 78% (74%, 81%) |
| **Immunocompromised** |  |  |  |  |  |  |  |  |  |
| Observed COVID-19 | 128 | 7,162 | 17.87 | 942 | 24,883 | 37.86 | 0.48 (0.40, 0.58) | 52% (42%, 60%) | 64% (57%, 70%) |
| COVID-19-related hospitalization | 39 | 7,177 | 5.43 | 301 | 24,994 | 12.04 | 0.46 (0.33, 0.65) | 54% (35%, 67%) | 68% (54%, 77%) |
| **Not immunocompromised** |  |  |  |  |  |  |  |  |  |
| Observed COVID-19 | 1,104 | 95,702 | 11.54 | 12,584 | 316,542 | 39.75 | 0.30 (0.28, 0.32) | 70% (68%, 72%) | 79% (78%, 81%) |
| COVID-19-related hospitalization | 204 | 95,846 | 2.13 | 2,809 | 318,058 | 8.83 | 0.25 (0.22, 0.29) | 75% (71%, 78%) | 83% (80%, 85%) |
| **High-Delta-incidence States\*\*** |  |  |  |  |  |  |  |  |  |
| Observed COVID-19 | 149 | 7,668 | 19.43 | 1,587 | 25,832 | 61.44 | 0.31 (0.26, 0.37) | 69% (63%, 74%) | 79% (75%, 83%) |
| June - July 2021 only*** | 110 | 4,491 | 24.49 | 1,084 | 14,814 | 73.17 | 0.33 (0.27, 0.40) | 67% (60%, 73%) | 78% (73%, 82%) |
| COVID-19-related hospitalization | 23 | 7,680 | 2.99 | 306 | 25,967 | 11.78 | 0.26 (0.17, 0.39) | 74% (61%, 83%) | 83% (74%, 89%) |
| June - July 2021 only*** | 12 | 4,500 | 2.67 | 171 | 14,918 | 11.46 | 0.23 (0.13, 0.41) | 77% (59%, 87%) | 85% (73%, 91%) |

Unless otherwise noted, incident rates are reported per 1,000 person-years and vaccine effectiveness (VE) estimates (observed and corrected) are calculated using hazard ratios with reported 95% confidence interval limits. Corrected VE estimates are adjusted for under-recording of vaccinations in claims data using the approach described in **Suppl. S4**.
\*\* High-Delta-incidence States (early Delta states) include Arkansas, Florida, Louisiana, and Missouri.
\*\*\* For June and July 2021 results within four states (Arkansas, Florida, Louisiana, and Missouri) with high prevalence of the Delta variant of concern, incident rate ratios (IRR) after PS matching are reported instead of hazard ratios and VE is estimated using $(1 - \text{IRR}) \times 100$ for patients contributing follow-up time from June 1, 2021 through July 31, 2021.

In high-Delta-incidence states, rates of observed COVID-19 were higher in both groups than in the national cohort, with incidence of 19.4/1,000 PY (vaccinated) v. 61.4/1,000 PY (unvaccinated) (**Table 2**). In these states, VE for observed COVID-19 was 79% (75% to 83%) overall and 78% (73% to 82%) during June and July (**Table 2**), the months where Delta variant incidence was highest; for COVID-19-related hospitalization VE was 83% (74% to 89%) overall and 85% (73% to 91%) during June and July. VE within subgroups of the high-Delta-incidence states for both outcomes were similar to the national cohort with VE for COVID-19 of 81% in patients <50 years of age and 84% for COVID-related hospitalization in patients >50 years (**Table S3**).

Examining the incidence of observed COVID-19 and COVID-19-related hospitalization as a function of time since vaccination, we observed sustained and stable VE starting 14 days after vaccination to a maximum of 152 days after vaccination (**Figure 2**). Inspection of Schoenfeld residual plots indicate that hazard ratios were constant over time and that there was no VE reduction during the observable follow-up time (**Figure S2**).

**Figure 2.**
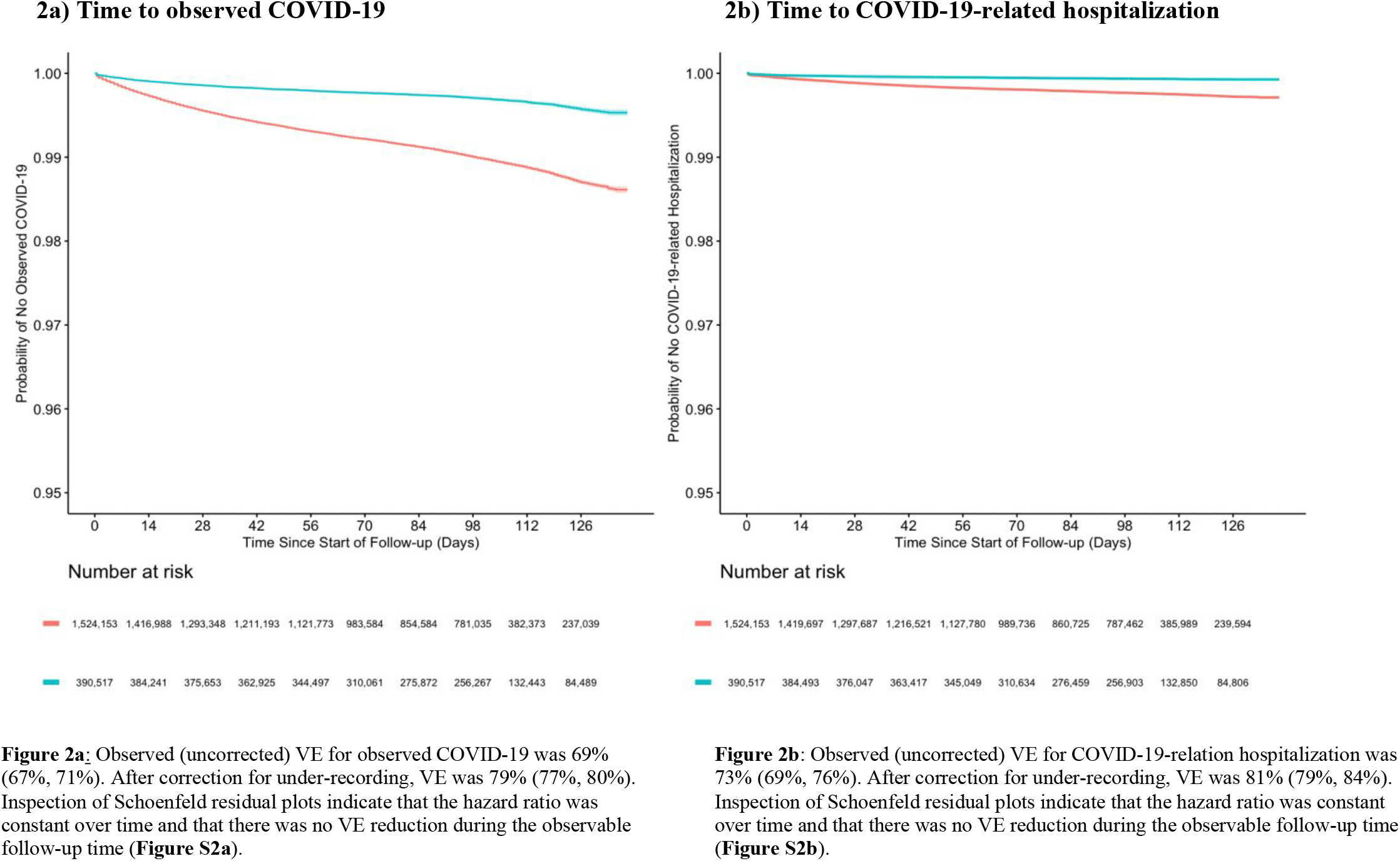
COVID-19-related outcomes by time since vaccination (+14 days) in national cohort. **Figures 2a and 2b**: Follow-up day 0 is equivalent to 14 days post-index, as follow-up for outcomes begins 14 days after vaccination or matched index date. Kaplan-Meier plots include 95% confidence intervals around the survival function.

An analysis by calendar months from March 2021 through July 2021 showed no meaningful variation in VE over calendar time. The VE for observed COVID-19 rose slightly until May to 81% (79% to 83%) and remained at a high level until the end of the follow-up period in July (77%; 74% to 79%) when the Delta variant was spreading widely nationally (**Figure 3a**). The monthly VE estimates for COVID-19-related hospitalization were equally stable (**Figure 3b**). In the high-Delta-incidence states we saw no decline in the VE with respect to hospitalizations during the peak Delta variant months of June and July (**Figure 3d)** and some more variability in the VE for observed COVID-19 **(Figure 3c**).

**Figure 3.**
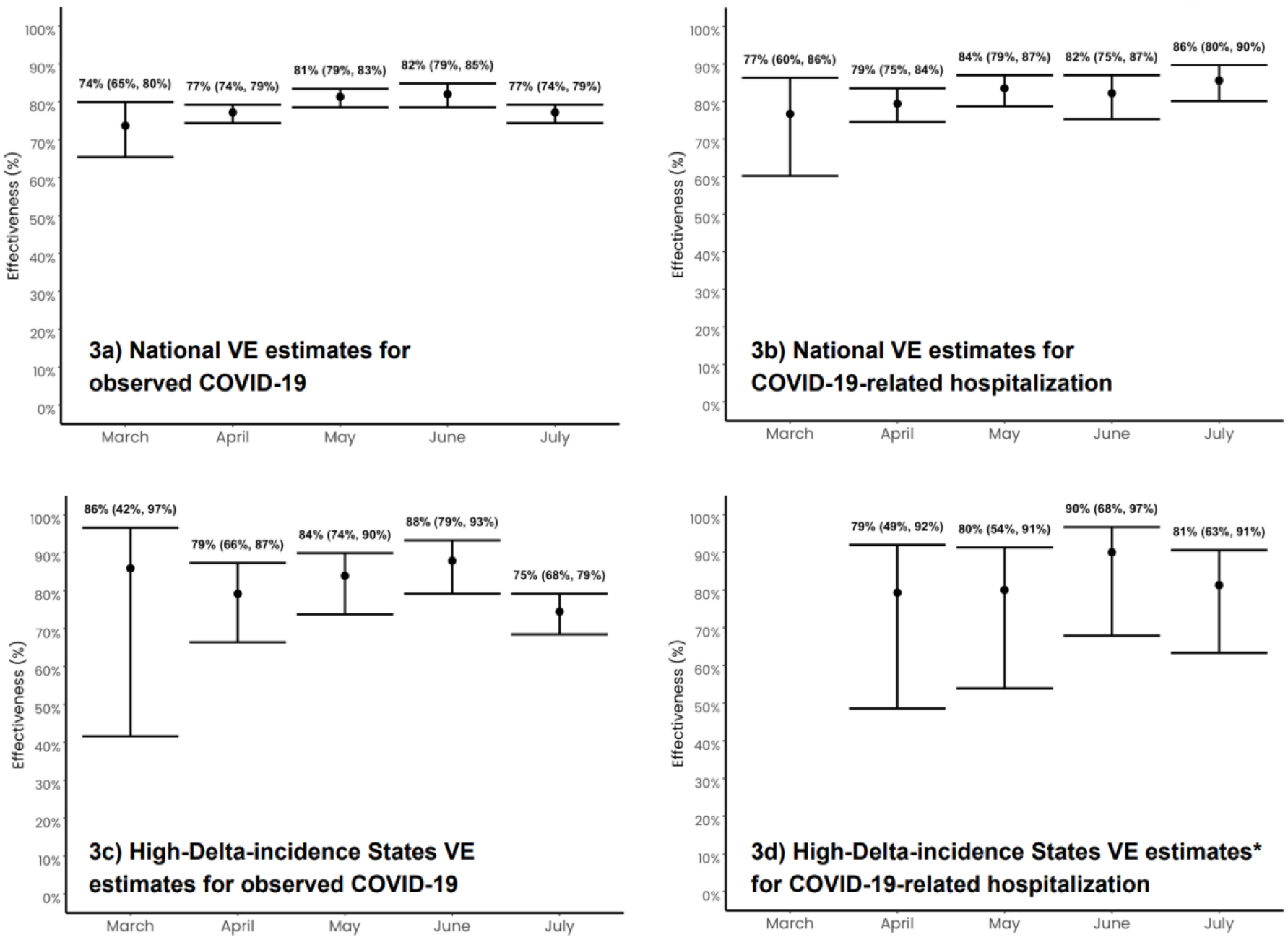
Vaccine effectiveness by month (March 2021 – July 2021) See **Suppl. S5** and **Figure S3** for further detail on methods used to calculate monthly VE estimates. All vaccine effectiveness estimates are corrected for under-recording (see **Suppl. S4**). * Insufficient outcome count for March-specific effectiveness calculations in Figure 3d.

Results in a sensitivity analysis defining COVID-19 using nucleic acid amplification test results were similar to primary findings (**Table S7**). When we required all participants to meet the additional criterion of COVID-related utilization before cohort entry, the corrected VE was reduced to 56% (53%, 60%) for COVID-19 and 57% (49%, 64%) for COVID-19-related hospitalization (**Table S8**).

## Discussion

In U.S. clinical practice from March through July 2021, the single-dose Ad26.COV2.S vaccine showed effective protection against observed COVID-19 and COVID-19-related hospitalization with no evidence of reduced effectiveness over the maximum study duration. In states with early emergence and high incidence of the Delta variant, we found equally high and sustained vaccine effectiveness (VE). These results provide evidence that the VE observed in the ENSEMBLE trial translates into clinical practice, lasts over at least 152 days post-vaccination, and remains effective amid high Delta variant incidence. Although we saw high VE in the US through July 2021, when the delta variant was dominant per CDC sequence data, we did not have sequence-specific information in our real-world data, hence we do not know with certainty the delta variant-specific vaccine effectiveness. We plan to update this Ad26.COV2.S VE study with monthly data refreshes to expeditiously identify any changes in VE.^25^

VE varied by age subgroup, with patients <50 having a higher VE than those ≥50 years of age. This is consistent with the age-dependent variation in VE observed in the ENSEMBLE trial and with evidence on the immune response to the BNT162b2 vaccine by age.^3,26,27^ Results from our population-based study suggest that immunocompromised patients who are vaccinated with a single dose have slightly reduced yet still substantial protection against COVID-19 and COVID-19-related hospitalizations for at least 150 days.

Although lacking baseline randomization, vaccinated and unvaccinated individuals were exactly matched by age, sex, comorbid-risk, calendar date, and location, followed by additional PS-matching on 17 risk factors for COVID-19 severity. In addition, we excluded individuals with prior COVID to avoid misclassifying post-COVID symptoms as new infections. Collectively, these techniques reduced the possibility of residual confounding. Unlike RCTs, the timeliness at which these analyses can be performed in national data -- and updated as external circumstances change -- address an otherwise un-addressable public health need. Further, our national and regional view of the data allow for characterization of VE in clinical practice, taking into account changing prevalence of variants, variations in government mandates, individual behavior, and other quickly-evolving factors.

This study is mainly based on the data that flows from U.S. healthcare providers to insurance companies for reimbursement and are available sooner than traditional adjudicated (“closed”) insurance claims. While other studies have used these “open” claims data to study COVID-19,^10^ the data (a) include no specific listing of who is enrolled in an insurance plan, making the denominator challenging to precisely estimate at a given point in time and (b) potentially lack complete capture of all longitudinal healthcare interactions for any given patient. Our study therefore required the occurrence of provider-based medical and pharmacy claims unrelated to COVID during baseline to restrict our population to those with more complete outcome capture, making it unlikely that there is differential capture of information between the groups. Further, the lack of meaningful month-to-month changes in VE demonstrating sustained protection of the vaccine is unlikely to be affected by this.

As a result of mass vaccination efforts by a range of public and private organizations, where health insurance information may not have been required, collected or submitted, a substantial fraction of COVID-19 vaccine administrations were not recorded in insurance data.^28^ Given the under-recording of COVID-19 vaccinations in insurance claims data which leads to an underestimation of the true effect size, we empirically estimated the level of under-recording in our data source (**Suppl. S4**) and applied standard correction methods to all VE estimates.^29^ Even if one would falsely assume 0% under-recording and completely disregard the necessary correction, the VE estimates would decline by about 10 percentage points as illustrated in **Tables S5 and S6**. The resulting biased and underestimated VE of 69% (67% to 71%) for infections and 73% (69% to 76%) for hospitalizations demonstrates substantial vaccine protection.

Recent seroprevalence data indicate a meaningful proportion of undocumented infections.^30^ Assuming such outcome misclassification would be non-differential regarding the vaccination status, this would likely lead to an underestimation of the reported VE on observed COVID-19 but would not affect the VE for COVID-related hospitalizations.^29^ Additionally, COVID-19 outcomes captured with ICD-10 diagnosis codes in claims data may indicate more severe cases requiring physician consult as opposed to individuals with asymptomatic or mild to moderate disease who may be less likely to seek professional care and generate claims.^31,32^ While it is unclear how exactly the infection severity definition compares with definitions used in RCTs, we suggest comparing our findings with the RCT’s severe, or moderate to severe, definition.^2^ Consistency of VE results across sensitivity analyses with lab-only outcome definitions provides further evidence for the robustness of our primary results (**Table S7**).

Finally, VE estimates were directionally consistent but lower in magnitude than primary results in sensitivity analyses requiring all vaccinated and unvaccinated individuals to have evidence of COVID-related utilization before cohort entry (**Table S8**). While this sensitivity analysis may reduce potential selection bias introduced during the data extraction stage by picking sicker unvaccinated individuals, it may also include more individuals with undiagnosed COVID before cohort entry, and thus may together represent the lower bound of VE.

These non-randomized population-based data from clinical practice in the US demonstrate the high and stable effectiveness of Ad26.COV2.S across high-risk patient subgroups and in geographic areas of high Delta variant incidence. As the COVID-19 pandemic continues and variants evolve the observed effectiveness may change in the future as new variants emerge. Our data and data to be generated over the coming months using this system of claims data contribute important and otherwise unattainable evidence to policymakers, physicians, and patients.

## Data Availability

Data belong to third parties and cannot be shared publicly, however, upon reasonable request, researchers may get access to the data and analytics infrastructure for pre-specified collaborative analyses.

## Acknowledgement

The authors thank the following team members for critical feedback during the development of the study protocol and/or review of findings: Deb Ricci (Janssen), Khaled Sarsour (Janssen), Jennifer Hayden (Janssen), Xiaoying Wu (Janssen), Sophia Gallucci (Aetion), Jen Thorburn (Aetion), Michelle Skornicki (Aetion), Rouhi Abdollahi (Janssen), Kristopher Standish (Janssen), Lev Demirdjian (Janssen), Daniel Backenroth (Janssen), and Jerry Sadoff (Janssen). We would like to also thank Andrew Kress (HealthVerity) for his support with technical data questions, and the many people involved in the technical implementation process and independent quality control, Julie Schiffman (Aetion), Hanaya Raad (Aetion), Julia Pitino (Aetion), Lexie Rubens (Aetion), and Tancy Zhang (Aetion).

