## Supplemental Methods and Figures for "Effectiveness of the Single-Dose Ad26.COV2.S COVID Vaccine"

**Supplement Contents**


| | |
|---|---|
| **<u>Supplementary Methods</u>** | |
| **Supplementary Methods 1 (Suppl. S1)** | Risk-set sampling of unvaccinated study individuals |
| **Supplementary Methods 2 (Suppl. S2)** | Codes used to identify cohort inclusion and exclusion variables |
| **Supplementary Methods 3 (Suppl. S3)** | Codes of pre-exposure covariates used for confounding adjustment via propensity score matching |
| **Supplementary Methods 4 (Suppl. S4)** | Vaccine under-reporting and correction |
| **Supplementary Methods 5 (Suppl. S5)** | Month-by-month effectiveness analyses |
| **<u>Supplementary Figures</u>** | |
| **Figure S1** | Study design diagram |
| **Figure S2** | Plot of Schoenfeld residuals for primary analysis in national cohort |
| **Figure S3** | Diagrams for methods used to calculate month-specific effectiveness estimates. |
| **<u>Supplementary Tables</u> (see attached excel document for supplementary tables)** | |
| **Table S0** | Study population counts and attrition |
| **Table S1** | Characteristics of Ad26.COV2.S vaccinated and matched unvaccinated individuals in national cohort, by age and immunocompromised subgroups. |
| **Table S2** | Characteristics of Ad26.COV2.S vaccinated and matched unvaccinated individuals in high-Delta-incidence States, overall and by age and immunocompromised subgroups. |
| **Table S3** | Incidence and vaccine effectiveness for COVID-19 and COVID-19-related hospitalizations - high-Delta-incidence States, overall and by age and immunocompromised subgroups. |
| **Table S4** | Incidence and vaccine effectiveness for COVID-19 and COVID-19-related hospitalizations - Regional subgroups. |
| **Table S5** | Correction of the VE regarding observed COVID-19 with varying assumptions about the level of under-recording in claims data. |

| | |
|---|---|
| **Table S6** | Correction of the VE regarding COVID-19-related hospitalizations with varying assumptions about the level of under-recording in claims data. |
| **Table S7** | Incidence and vaccine effectiveness for COVID-19 and COVID-19-related hospitalizations defined by laboratory NAAT test results only. |
| **Table S8** | Incidence and vaccine effectiveness for COVID-19 and COVID-19-related hospitalizations among cohort meeting data extraction criteria before cohort entry. |
| **Table S9** | Characteristics of Ad26.COV2.S vaccinated and risk-set sample matched unvaccinated individuals in national cohort prior to propensity score matching. |

**Supplementary Methods 1 (Suppl. S1): Risk-set sampling of unvaccinated study individuals**
A two-step process was used to identify a referent group of non-vaccinated individuals available for follow-up on the same date and geography as all identified exposed individuals. This two-step process first included risk-set sampling via exact matching exposed (vaccinated) individuals with up to ten non-vaccinated (referent) individuals on: time (calendar date), age (within age categories of 18-24; 25-29; 30-34; 35-39; 40-44; 45-49; 50-54; 55-59; 60-64; 65-69; 70-74; 75-79; 80-84; 85+), sex, geography (based on 3-digit ZIP codes), and Gagne Combined Comorbidity Index based on 365 day baseline period (categories: 0-1, 2-3, 4-5, 6+).

From these risk-set sampled patients, PS matching as described in the manuscript was performed to obtain final matched cohorts.

**Supplementary Methods 2 (Suppl. S2): Codes used to identify cohort inclusion and exclusion variables**

| **Variable name** | **RWD Definition** |
|---|---|
| Administration of the Janssen Ad26.COV2.S vaccine (inclusion) | Any medical claim, pharmacy claim, inpatient hospital encounter, or outpatient hospital encounter with one of the following CPT or NDC codes recorded:<br>CPT codes: 91303, 0031A<br>NDC codes: 59676-580-05, 59676-0580-05, 59676-0580-15, 59676-580-15<br>OR<br>Any retail pharmacy claim for a COVID-19 vaccine with the following manufacturer name: “JANSSEN”, “JANSSEN (DIVISION OF JOHNSON AND JOHNSON)”, “JOHNSON AND JOHNSON”, “JSN” and CVX code is 212<br><br>Note: CVX codes are codes that indicate the product used in a vaccination and are maintained by the CDC’s National Center of Immunization and Respiratory Diseases. |
| Evidence of prior observed COVID-19 (exclusion) | Any medical claim, inpatient hospital encounter, or outpatient hospital encounter with one of the following ICD-10-CM codes:<br>U07.1<br>Z20.822<br>Z86.16 |

| | |
|---|---|
| | J12.82<br>OR<br>Record of any positive result from a COVID-19 diagnostic or antibody laboratory test. |
| Administration of any COVID-19 vaccine (exclusion) | Administration of the Janssen Ad26.COV2.S vaccine<br>OR<br>Administration of an mRNA vaccine (Pfizer or Moderna):<br>{Any medical claim, pharmacy claim, inpatient hospital encounter, or outpatient hospital encounter with one of the following CPT or NDC codes recorded:<br>CPT codes: 0001A, 0002A, 0012A, 0011A, 91300, 91301<br>NDC codes: 80777-0273-10, 59267-1000-01, 59267-1000-1, 80777-273-10, 59267-1000-2, 59267-1000-02, 59267-1000-03, 80777-0273-15, 80777-0273-98, 80777-273-15, 80777-273-99, 80777-0273-99<br>OR<br>Any retail pharmacy claim for a COVID-19 vaccine with the following manufacturer name:<br>“MOD”, “MODERNA”, “MODERNA US, INC”, “MODERNA US, INC.”, “PFIZER”, “PFIZER, INC”, “PFR” and CVX code is 207 or 208<br><br>Note: CVX codes are codes that indicate the product used in a vaccination and are maintained by the CDC’s National Center of Immunization and Respiratory Diseases. |

**Supplementary Methods 3 (Suppl. S3): Codes of pre-exposure covariates used for confounding adjustment via propensity score matching**

| Variable name | RWD Definition |
|---|---|
| Chronic obstructive pulmonary disease (COPD) | Any medical claim, inpatient hospital encounter, or outpatient hospital encounter with one of the following ICD-10-CM codes[1]: I27.8, I27.9, J40, J41, J42, J43, J44, J45, J46, J47, J60, J61, J62, J63, J64, J65, J66, J67, J68.4, J70.1, J70.3 |
| Pulmonary fibrosis | Any medical claim, inpatient hospital encounter, or outpatient hospital encounter with one of the following ICD-10-CM codes[2]: J61, J62.0, J62.8, J63.0, J63.1, J63.2, J63.3, J63.4, J63.5, J63.6, J64, J65, J66.0, J66.1, J66.2, J66.8, J70.1, J84.01, J84.02, J84.03, J84.09, J84.10, J84.111, J84.112, J84.113, J84.114, J84.115, J84.116, J84.117, J84.170, J84.178, J84.2, J84.841, J84.842, J84.843, J84.848, J84.89, J84.9 |
| HIV infection | Any medical claim, inpatient hospital encounter, or outpatient hospital encounter with one of the following ICD-10-CM codes[3]: B20, B97.35, R75, Z21 |
| Immunocompromised status (as used for subgroups) | Moderately to severely immunocompromised subgroups were defined as those with any diagnosis for active cancer, history of organ/stem cell transplant, primary immunodeficiency (e.g., DiGeorge syndrome, Wiskott-Aldrich syndrome), or HIV infection in the 365d baseline period,<br>AND/OR<br>Recent use (within 60 days of index) of immunosuppressive therapies including high-dose corticosteroids (i.e., ≥20mg prednisone or equivalent per day), transplant-related immunosuppressives, antimetabolites, alkylating agents, and other severely immunosuppressive cancer chemotherapeutics, tumor necrosis factor blockers, and other immunosuppressive biologics. Immunocompromised subgroup definitions based on CDC vaccine guidance for moderately to severely immunocompromised status.<br><br>**Operationalized in claims data as:**<br>Occurrence of Immunocompromised state from blood transplant during 365d baseline period<br>OR<br>Occurrence of Immunocompromised state from organ transplant during 365d baseline period<br>OR<br>Occurrence of HIV infection during 365d baseline period<br>OR<br>Any medical claim, inpatient hospital encounter, or outpatient hospital encounter with one of the following ICD-10-CM codes during 365d baseline period: B20, B97.35, D45, D46.22, D47.1, D47.4, D47.9, D47.Z1, D47.Z9, D61.82, D75.81, D82, D82.0, D82.1, D82.2, D82.3, D82.4, D82.8, D82.9, R75, Z21, Diagnosis for active malignancy among C00-C96 (excluding sub-codes for neoplasms, benign tumors)<br>OR<br>Any medical claim, inpatient hospital encounter, or outpatient hospital encounter with one of the following CPT/HCPCS procedure codes for organ transplantation and related during 365d baseline period: 32852, 32853, 32854, 3490F, 38243, 50365, 50370, A4653, A4671, A4673, A4690, A4700, A4707, A4709, A4712, A4714, A4722, A4725, A4726, A4730, A4737, A4740, A4750, A4755, A4765, A4860, A4890, A4910, A4912, A4913, A4918, E1500, E1510, E1520, E1530, E1540, E1550, E1590, E1592, E1600, E1625, E1634, E1635, S2053, S2054, |

| | |
|---|---|
| | S2060, S2152, 32851, 33935, 33945, 38240, 38242, 44135, 44136, 47135, 47136, 48554, 50360, A4672, A4674, A4680, A4705, A4706, A4708, A4719, A4720, A4721, A4723, A4724, A4728, A4735, A4736, A4760, A4766, A4802, A4820, A4850, A4870, A4880, A4900, A4901, A4905, A4911, A4914, E1560, E1570, E1575, E1580, E1594, E1610, E1615, E1620, E1630, E1632, E1636, S2065, S2142, S2150<br>OR<br>Any medical claim, inpatient hospital encounter, or outpatient hospital encounter with one of the following ICD-10-PCS procedure codes for organ transplantation and related during the 365d baseline period: 02YA0Z2, 07YM0Z2, 07YP0Z0, 07YP0Z1, 07YP0Z2, 0BYC0Z2, 0BYD0Z1, 0BYD0Z2, 0BYF0Z0, 0BYG0Z1, 0BYG0Z2, 0BYJ0Z0, 0BYJ0Z2, 0BYK0Z2, 0BYL0Z2, 0BYM0Z1, 0BYM0Z2, 0DY50Z0, 0DY50Z1, 0DY50Z2, 0DY60Z2, 0DY80Z0, 0DYE0Z1, 0FY00Z1, 0FY00Z2, 0FYG0Z1, 0FYG0Z2, 0TY00Z0, 0TY00Z2, 0TY10Z1, 0TY10Z2, 30230G1, 30230G2, 30230G4, 30230X3, 30230X4, 30230Y1, 30230Y3, 30233G1, 30233G4, 30233X1, 30233X4, 30233Y1, 30233Y4, 30240AZ, 30240G4, 30240X1, 30240X2, 30240X4, 30240Y1, 30240Y2, 30243AZ, 30243G2, 30243G3, 30243G4, 30243X1, 30243X4, 30243Y2, 30243Y4, 30250G1, 30253G1, 30253X1, 30260G1, 30260X1, 5A1D00Z, 5A1D80Z, 5A1D90Z, BT2900Z, BT290ZZ, BT29Y0Z, BT29YZZ, BT29ZZZ, BT39YZZ, BT39ZZZ, 02YA0Z0, 02YA0Z1, 07YM0Z0, 07YM0Z1, 0BYC0Z0, 0BYC0Z1, 0BYD0Z0, 0BYF0Z1, 0BYF0Z2, 0BYG0Z0, 0BYH0Z0, 0BYH0Z1, 0BYH0Z2, 0BYJ0Z1, 0BYK0Z0, 0BYK0Z1, 0BYL0Z0, 0BYL0Z1, 0BYM0Z0, 0DY60Z0, 0DY60Z1, 0DY80Z1, 0DY80Z2, 0DYE0Z0, 0DYE0Z2, 0FY00Z0, 0FYG0Z0, 0TY00Z1, 0TY10Z0, 30230AZ, 30230G3, 30230X1, 30230X2, 30230Y2, 30230Y4, 30233AZ, 30233G2, 30233G3, 30233X2, 30233X3, 30233Y2, 30233Y3, 30240G1, 30240G2, 30240G3, 30240X3, 30240Y0, 30240Y3, 30240Y4, 30243G1, 30243X2, 30243X3, 30243Y1, 30243Y3, 30250X1, 30250Y1, 30253Y1, 30260Y1, 30263G1, 30263X1, 30263Y1, 3E1M39Z, 5A1D70Z, BT2910Z, BT291ZZ, BT39Y0Z, BT49ZZZ<br>OR<br>Any medical claim, pharmacy claim, inpatient hospital encounter, or outpatient hospital encounter with one of the following NDC Generic Names recorded during the 365d baseline period:<br>azacitidine, bendamustine hcl, busulfan, capecitabine, carmustine, carmustine in polifeprosan 20, chlorambucil, clofarabine, cyclophosphamide, cytarabine, cytarabine liposome/pf, cytarabine/pf, dacarbazine, daunorubicin/cytarabine liposomal, decitabine, decitabine/cedazuridine, diroximel fumarate, floxuridine, fludarabine phosphate, fluorouracil, fluorouracil/adhesive bandage, gemcitabine hcl, gemcitabine hcl in 0.9 % sodium chloride, ifosfamide, ifosfamide/mesna, inebilizumab-cdon, lomustine, melphalan, melphalan hcl, melphalan hcl/betadex sulfobutyl ether sodium, mercaptopurine, nelarabine, ofatumumab, ozanimod hydrochloride, pemetrexed disodium, pralatrexate, satralizumab-mwge, streptozocin, temozolomide, teprotumumab-trbw, thiotepa, upadacitinib, baricitinib, canakinumab/pf, cyclosporine/chondroitin sulfate a sodium, efalizumab, emapalumab-lzsg, guselkumab, infliximab-abda, infliximab-axxq, muromonab-cd3, natalizumab, ocrelizumab, ravulizumab-cwvz, risankizumab-rzaa, sarilumab, siltuximab, tacrolimus/niacinamide, tildrakizumab-asmn, vedolizumab, anakinra, belatacept, certolizumab pegol, daclizumab, eculizumab, infliximab, infliximab-dyyb, rilonacept, tacrolimus in vehicle base no.238, tacrolimus, micronized, tacrolimus/hyaluronate sodium/niacinamide, ustekinumab, abatacept/maltose, azathioprine sodium, pirfenidone, pomalidomide, secukinumab, teriflunomide, abatacept, alefacept, alemtuzumab, basiliximab, brodalumab, golimumab, mycophenolate mofetil hcl, temsirolimus, tofacitinib citrate, belimumab, siponimod, tacrolimus anhydrous, apremilast, thalidomide, fingolimod hcl, ixekizumab, tocilizumab, dimethyl fumarate, lenalidomide, cladribine, etanercept, adalimumab, methotrexate, mycophenolate sodium, methotrexate/pf, cyclosporine, |

| | |
|---|---|
| | modified, everolimus, leflunomide, sirolimus, azathioprine, cyclosporine, mycophenolate mofetil, methotrexate sodium/pf, methotrexate sodium, tacrolimus<br>OR<br>Any medical claim, pharmacy claim, inpatient hospital encounter, or outpatient hospital encounter with one of the following CPT/HCPC procedure codes recorded during the 365d baseline period:<br>80158, 80197, C9006, C9020, C9026, C9106, C9110, C9211, C9212, C9230, C9236, C9239, C9249, C9261, C9421, C9455, J0129, J0135, J0202, J0480, J0490, J0638, J1300, J1602, J1628, J2323, J2793, J3245, J3357, J7504, J7507, J7508, J7513, J7515, J7517, J7520, J8530, J9092, J9094, J9095, J9250, J9260, J9310, J9312, J9330, K0120, K0121, K0412, Q2019, Q2044, Q4079, Q5109, S0087, S0162, S0193, S9359, 80169, 80180, 80195, C9126, C9219, C9264, C9286, C9419, C9420, C9436, C9438, J0215, J0485, J0717, J0718, J1438, J1745, J2350, J2860, J3262, J3358, J3380, J7500, J7501, J7502, J7503, J7505, J7511, J7518, J7525, J7527, J8561, J8610, J9010, J9065, J9070, J9080, J9090, J9091, J9093, J9096, J9097, J9311, K0119, K0122, K0123, Q5103, Q5104<br>**Reference and evidence for association:**<br>Immunocompromised subgroup definitions based on CDC vaccine guidance for moderately to severely immunocompromised status. |
| Immunocompromised state from blood transplant | Any medical claim, inpatient hospital encounter, or outpatient hospital encounter with one of the following ICD-10-CM procedure codes: 30230AZ, 30230G1-G4, 30230X1-X4, 30230Y1-Y4, 30233AZ, 30233G1-G4, 30233X1-S4, 30233Y1-Y4, 30240AZ, 30240G1-G4, 30240X1-X4, 30240Y1-Y4, 30243AZ, 30243G1-G4, 30243X1-X4, 30243Y1-Y4, 30250G1, 30250X1, 30250Y1, 30253G1, 30253X1, 30253Y1, 30260G1, 30260X1, 30260Y1, 30263G1, 30263X1, 30263Y1<br>OR<br>ICD-10 diagnosis codes, D84.81, D84.9, D81.1, T86.00, T86.09, Z48.290, Z94.81 |
| Immunocompromised state from organ transplant | Any medical claim, inpatient hospital encounter, or outpatient hospital encounter with one of the following codes[4]:<br>DRG procedure codes: kidney (302), heart (103), lung (495), liver (480)<br>OR<br>ICD-10 codes: D84.821, D84.9, Z48.21, Z48.22, Z48.23, Z48.24, Z48.280, Z48.288, Z94.0, Z94.1, Z94.2, Z94.3, Z94.4, Z94.82, Z94.83, T86.10, T86.19, T86.20, T86.298, T86.30, T86.39, T86.40, T86.49, T86.818, T86.819, T86.858, T86.859, T86.898, T86.899 |
| Liver disease | Any medical claim, inpatient hospital encounter, or outpatient hospital encounter with one of the following ICD-10-CM codes[1]: B15.9, B16.0, B16.1, B16.2, B16.9, B17.0, B17.10, B17.11, B17.2, B17.8, B17.9, B18, B19.0, B19.10, B19.11, B19.20, B19.21, B19.9, B66.1, B66.3, I85, I86.4, I98.2, K70, K71.0, K71.1, K71.10, K71.11, K71.2, K71.3, K71.4, K71.5, K71.50, K71.51, K71.6, K71.7, K71.8, K71.9, K72, K73, K74, K75.0, K75.1, K75.2, K75.3, K75.4, K75.81, K75.89, K75.9, K76.0, K76.1, K76.2, K76.3, K76.4, K76.5, K76.6, K76.7, K76.8, K76.81, K76.89, K76.9, K77, Q44.6, Z94.4 |
| Malignancies (excluding non-melanoma skin cancer) | Any medical claim, inpatient hospital encounter, or outpatient hospital encounter with one of the following ICD-10-CM codes[1]:<br>C00, C01, C02, C03, C04, C05, C06, C07, C08, C09, C10, C11, C12, C13, C14, C15, C16, C17, C18, C19, C20, C21, C22, C23, C24, C25, C26, C30, C31, C32, C33, C34, C37, C38, C39, C40, C41, C43, C45, C46, C47, C48, C49, C50, C51, C52, C53, C54, C55, C56, C57, |

| | |
|---|---|
| | C58, C60, C61, C62, C63, C64, C65, C66, C67, C68, C69, C70, C71, C72, C73, C74, C75, C76, C81, C82, C83, C84, C85, C88, C90, C91, C92, C93, C94, C95, C96, C97 |
| Moderate-to-severe asthma | Any medical claim, inpatient hospital encounter, or outpatient hospital encounter with one of the following ICD-10-CM codes[5]: J45.40, J45.41, J45.42, J45.50, J45.51, J45.52 |
| Cerebrovascular disease | Any medical claim, inpatient hospital encounter, or outpatient hospital encounter with one of the following ICD-10-CM codes[1]: G45, G46, H34.0, I60, I61, I62, I63, I64, I65, I66, I67, I68, I69 |
| Chronic kidney disease | Any medical claim, inpatient hospital encounter, or outpatient hospital encounter with one of the following ICD-10-CM codes[1]: I12.0, 13.1, N03.2, N03.3, N03.4, N03.5, N03.6, N03.7, N05.2, N05.3, N05.4, N05.5, N05.6, N05.7, N18, N19, N25.0, Z49.0, Z49.1, Z49.2, Z94.0, Z99.2 |
| Hypertension | Any medical claim, inpatient hospital encounter, or outpatient hospital encounter with one of the following ICD-10-CM codes[1]: I10, I11, I12, I13, I15, I16, I27.0, I27.1, I27.20, I27.21, I27.22, I27.23, I27.24, I27.29, I67.4, I87.301, I87.302, I87.303, I87.309, I87.311, I87.312, I87.313, I87.319, I87.31, I87.321, I87.322, I87.323, I87.329, I87.331, I87.332, I87.333, I87.339, I87.391, I87.392, I87.393, I87.399, I97.3, K76.6, R03.0, G93.2 |
| Serious heart conditions | Any medical claim, inpatient hospital encounter, or outpatient hospital encounter with one of the following ICD-10-CM codes[1,6]: 109.9, 111, 113, 113.2, I25.5, I42.0, I42.5-I42.9, I43, I50, P29.0, I20, I21, I22, I23, I24, I25, I05, I06, I07, I08, I20.0, I20.1, I20.8, I20.9, I23.7, I24.0, I24.1, I24.8, I24.9, I25.10, I25.110, I25.111, I25.118, I25.119, I25.2, I25.5, I26.01, I26.02, I26.09, I26.90, I26.92, I26.99, I27.0, I27.1, I27.2, I27.81, I27.82, 414.1, 414.11, 414.12, 414.19, 429.1, 429.2, 429.3, 429.5, 429.6, 429.71, 429.79, 429.81 |
| Obesity | Any medical claim, inpatient hospital encounter, or outpatient hospital encounter with one of the following ICD-10-CM codes[1,7]: Z68.30, Z68.31, Z68.32, Z68.33, Z68.34, Z68.35, Z68.36, Z68.37, Z68.38, Z68.39, Z68.41, Z68.42, Z68.43, Z68.44, Z68.45, Z68.51, Z68.52, Z68.53, Z68.54, Z98.0, Z98.84, E66 |
| Sickle Cell Disease | Any medical claim, inpatient hospital encounter, or outpatient hospital encounter with one of the following ICD-10-CM codes:<br>D57 |
| Thalassemia | Any medical claim, inpatient hospital encounter, or outpatient hospital encounter with one of the following ICD-10-CM codes:<br>D56 |
| Type 1 diabetes mellitus | Any medical claim, inpatient hospital encounter, or outpatient hospital encounter with one of the following ICD-10-CM codes[1]:<br>E10 |
| Type 2 diabetes mellitus | Any medical claim, inpatient hospital encounter, or outpatient hospital encounter with one of the following ICD-10-CM codes[1]:<br>E11 |

| | |
|---|---|
| Neurologic conditions | Any medical claim, inpatient hospital encounter, or outpatient hospital encounter with one of the following ICD-10-CM codes[3,8]: F01, F00, G00.0, G00.1, G00.2, G00.3, G00.8, G00.9, G01-G14, G20-G26, G30-G32, G31 (exclude G31.2), G35-37, G40, G43-G47, G50-G65, G70-G73, G80-G83, G89-99, G45.8, G45.9, I60-I63, I65-I69, F03, F04, R29, R41, S06, Z86.73, Z87.820, F02.80, F02.81, F05, F06.0, F06.8, F10.27, F19.97 |
| Number of pharmacy claims | The count of events occurring in the "Pharmacy Claims" table over the baseline period |
| Number of medical claims | The count of events occurring in the "Medical Claims" table over the baseline period |
| Recent medical claims | True if any event begins in the "Medical Claims" table in the 60 days prior to index |
| Recent pharmacy claim | True if any event begins in the "Pharmacy Claims" table in the 60 days prior to index |
| Age | Age defined as continuous numeric variable for propensity score modeling. |
| Sex | Male/Female |
| Comorbidity score (exact) | Gagne Combined Comorbidity Index |
| Calendar month of index date | The calendar month in which the index date occurs may be one of the following: 2021-03, 2021-04, 2021-05, 2021-06, 2021-07 |
| State | Classified based on U.S. state, D.C., or miscellaneous U.S. territory recorded on index date. |

**Supplementary Methods 4 (Suppl. S4): Vaccine under-recording and correction**

To assess under-recording of vaccination, we compared CDC-reported vaccination numbers to the number of observed COVID-19 vaccinations within the national open claims data used for this study. The CDC reported nationwide 162 million fully vaccinated people out of 283.7 million people 12 years and older in the US (~57%) as of July 22, 2021. Within the HealthVerity dataset used for the current study (data through July 2021), we observed 54 million individuals with any COVID-19 vaccination out of 161 million people included in the dataset (~34%). The under-recording of COVID-19 vaccinations in the national claims data used for this study can thus be approximated by 34%/57% ≈ 41% under-recording, which equates to ~59% sensitivity of exposure assessment.

Such reduced sensitivity of exposure assessment leads to an underestimation of the true effectiveness, as our unvaccinated comparator population likely includes patients who were vaccinated at mass vaccination sites, pharmacies, and other organizations without submitting claims to insurance companies and thus our study data source.[9] In order to be conservative, our primary analysis assumed an under-reporting of 40% (sensitivity of 60%) for all bias-corrected effect estimates.

Assuming a 40% estimated proportion of vaccination under-reporting in claims data, we used standard algebraic methods to correct relative risk estimates assuming 100% specificity of vaccination recording.[10] Observed and corrected relative risks and 95% confidence intervals were calculated by specifying a 61-day fixed follow-up period that began 14 days post-index among vaccinated and 1:1 PS matched unvaccinated patients. From this fixed follow-up analysis, specific bias correction factors were derived for each outcome[10] and all subgroups of interest, and then applied to hazard ratio and rate ratio-based effectiveness estimates from the primary analysis.

**Table S5. Correction of the VE regarding observed COVID-19 outcomes with varying assumptions about the level of under-recording in claims data**

| Assumed under-recording of COVID-19 vaccination status in national claims data | 0% | 10% | 20% | 30% | 40%* | 50% | 60% | 70% |
|---|---|---|---|---|---|---|---|---|
| Sensitivity of COVID-19 vaccine exposure | 100% | 90% | 80% | 70% | 60% | 50% | 40% | 30% |
| Uncorrected (observed) HR (95% CI) | 0.31 (0.29, 0.33) | 0.31 (0.29, 0.33) | 0.31 (0.29, 0.33) | 0.31 (0.29, 0.33) | 0.31 (0.29, 0.33) | 0.31 (0.29, 0.33) | 0.31 (0.29, 0.33) | 0.31 (0.29, 0.33) |
| Uncorrected (observed) Effectiveness (95% CI) | 69% (67%, 71%) | 69% (67%, 71%) | 69% (67%, 71%) | 69% (67%, 71%) | 69% (67%, 71%) | 69% (67%, 71%) | 69% (67%, 71%) | 69% (67%, 71%) |
| Corrected HR (95% CI) | 0.31 (0.29, 0.33) | 0.29 (0.27, 0.31) | 0.27 (0.25, 0.28) | 0.24 (0.23, 0.26) | 0.21 (0.20, 0.23) | 0.19 (0.17, 0.20) | 0.16 (0.15, 0.17) | 0.12 (0.11, 0.13) |
| Corrected Effectiveness (95% CI) | 69% (67%, 71%) | 71% (69%, 73%) | 73% (72%, 75%) | 76% (74%, 77%) | 79% (77%, 80%) | 81% (80%, 83%) | 84% (83%, 85%) | 88% (87%, 89%) |

* Primary Analysis

**Table S6. Correction of the VE regarding COVID-19-related hospitalizations with varying assumptions about the level of under-recording in claims data**

| Assumed under-recording of COVID-19 vaccination status in national claims data | 0% | 10% | 20% | 30% | 40%* | 50% | 60% | 70% |
|---|---|---|---|---|---|---|---|---|
| Sensitivity of COVID-19 vaccine exposure | 100% | 90% | 80% | 70% | 60% | 50% | 40% | 30% |
| Uncorrected (observed) HR (95% CI) | 0.31 (0.29, 0.33) | 0.31 (0.29, 0.33) | 0.31 (0.29, 0.33) | 0.31 (0.29, 0.33) | 0.31 (0.29, 0.33) | 0.31 (0.29, 0.33) | 0.31 (0.29, 0.33) | 0.31 (0.29, 0.33) |
| Uncorrected (observed) Effectiveness (95% CI) | 73% (69%, 76%) | 73% (69%, 76%) | 73% (69%, 76%) | 73% (69%, 76%) | 73% (69%, 76%) | 73% (69%, 76%) | 73% (69%, 76%) | 73% (69%, 76%) |
| Corrected HR (95% CI) | 0.27 (0.24, 0.31) | 0.25 (0.22, 0.29) | 0.23 (0.20, 0.26) | 0.21 (0.19, 0.24) | 0.19 (0.16, 0.21) | 0.16 (0.14, 0.18) | 0.13 (0.12, 0.15) | 0.10 (0.09, 0.12) |
| Corrected Effectiveness (95% CI) | 73% (69%, 76%) | 75% (71%, 78%) | 77% (74%, 80%) | 79% (76%, 81%) | 81% (79%, 84%) | 84% (82%, 86%) | 87% (85%, 88%) | 90% (88%, 91%) |

* Primary Analysis

**Supplementary Methods 5 (Suppl. S5): Month-by-month effectiveness analyses**

To assess vaccine effectiveness over calendar time, we examined incident rate ratios separately for each calendar month within the overall national cohort and high-Delta-incidence States (**Fig 3**). **Table 2** also shows effectiveness estimates for the combined period June to July 2021 for high-Delta-incidence states, given the emergence of the Delta variant during this two-month period.

Only outcome events and eligible person-time occurring within each person's follow-up period for main effectiveness analyses (March 2021 to July 2021) were included in month-level incident rate ratio calculations. Persons could contribute eligible person-time for multiple months, and/or for partial months, but were excluded from monthly person-time totals if censored from analysis. **Figure S3** shows how month-by-month person-time denominators were derived for 2 hypothetical study participants.

Using this approach, monthly incident rates were calculated as the (number of outcome events beginning in each calendar month) / (the total number of person-years within each month for which persons were eligible for outcomes). Incident rate ratios were calculated for each month as the ratio of outcome rates in exposed vs unexposed patients, and effectiveness estimates were then corrected for vaccine under-recording in claims data using the methods described in **Suppl. S4.**

**Figure S1. Study design diagram**

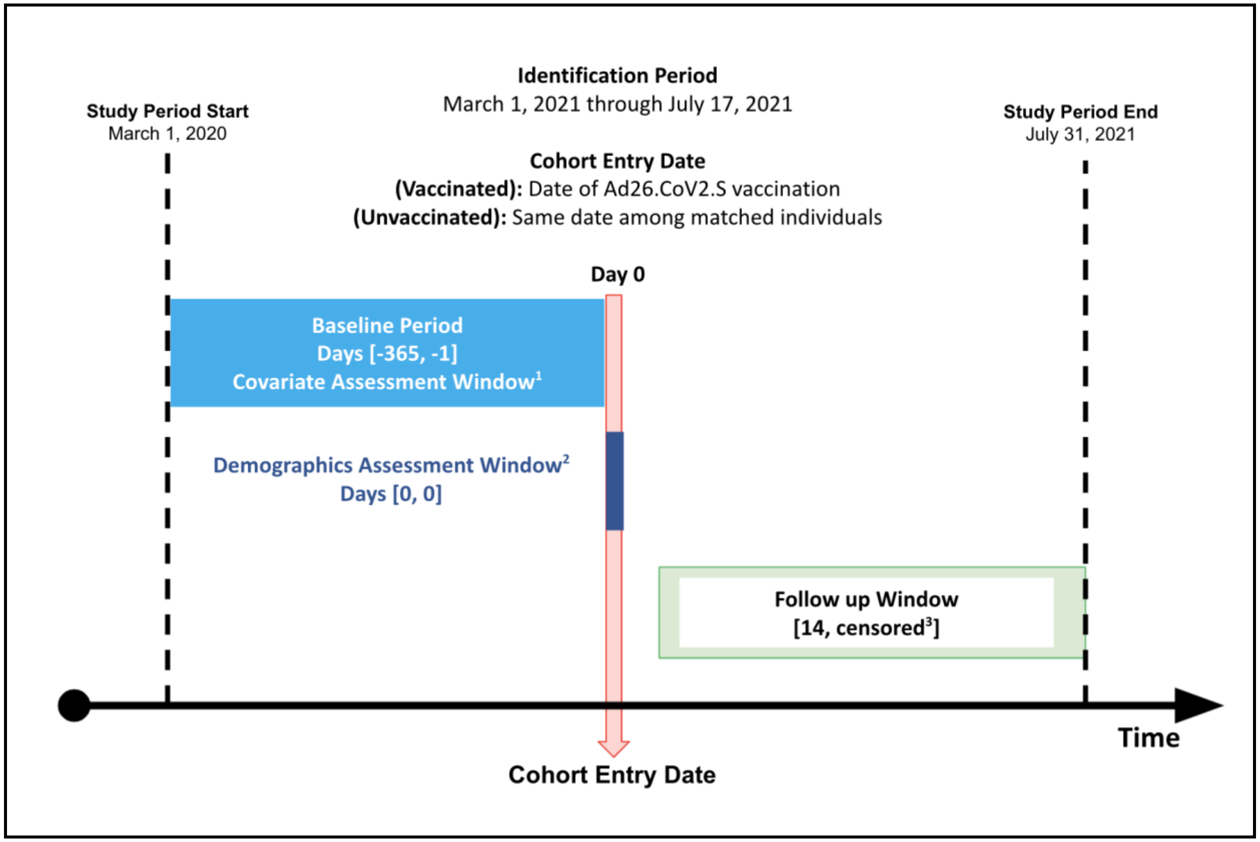


1 - Number of pharmacy/medical claims and comorbidities

2 - Age, sex and state

3 - Day 14 through observed endpoint or censoring

**Figure S2. Plot of Schoenfeld residuals for primary analysis in national cohort**

**S2a) Plot of Schoenfeld residuals for any observed COVID-19**

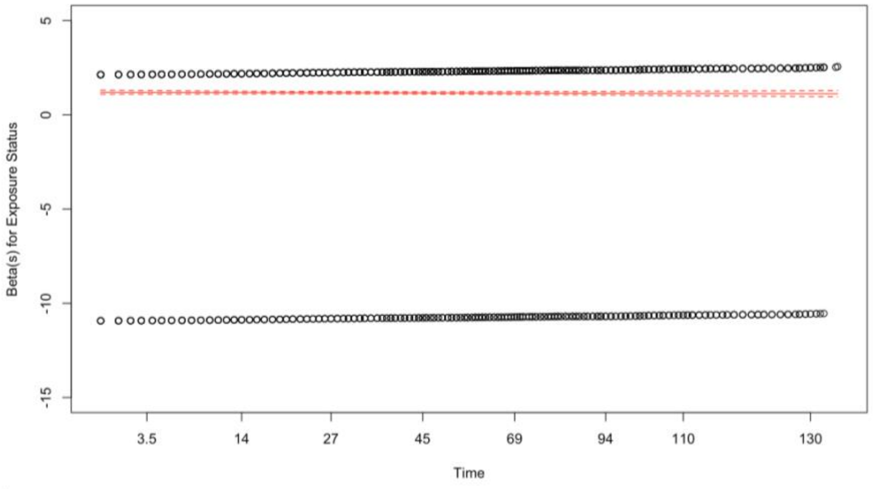


**S2b) Plot of Schoenfeld residuals for COVID-19-related hospitalization**

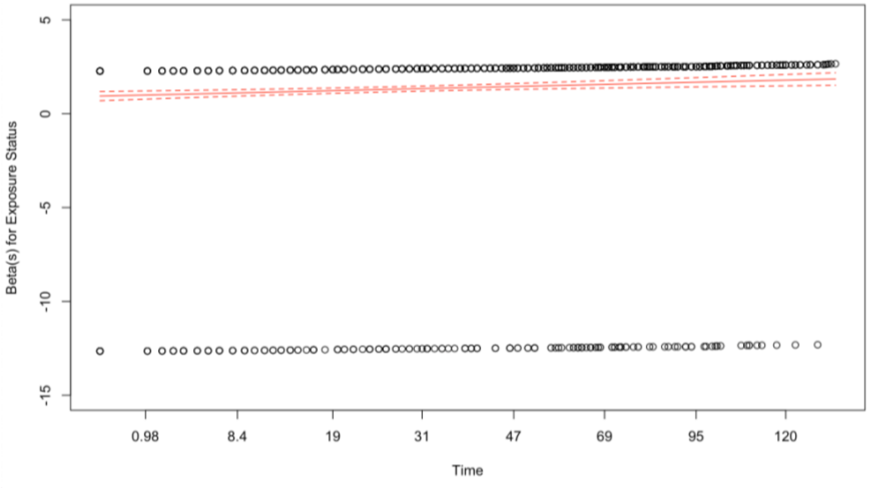

**Figure S3. Diagrams for methods used to calculate month-specific effectiveness estimates**

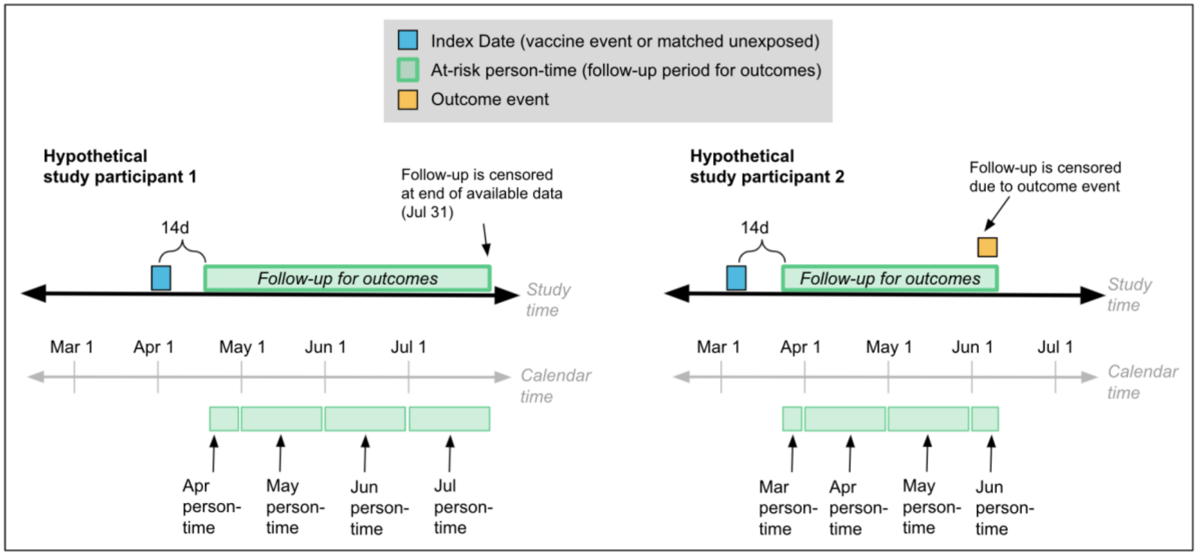


***Hypothetical study participant 1** contributes to separate person-time denominators for part of April, May, June, and July. Person 1 has no outcome events during this eligible time window.*

***Hypothetical study participant 2** contributes to separate person-time denominators for part of March, April, May, and part of June, and an outcome event in June.*
